# Lower respiratory tract myeloid cells harbor SARS-CoV-2 and display an inflammatory phenotype

**DOI:** 10.1101/2020.08.11.20171967

**Authors:** William Bain, Hernán F. Peñaloza, Mark S. Ladinsky, Rick van der Geest, Mara Sullivan, Mark Ross, Georgios D. Kitsios, Barbara Methe, Bryan J. McVerry, Alison Morris, Alan M. Watson, Simon C. Watkins, Claudette M. St Croix, Donna B. Stolz, Pamela J. Bjorkman, Janet S. Lee

## Abstract

SARS-CoV-2 pneumonia may induce an aberrant immune response with brisk recruitment of myeloid cells into the lower respiratory tract, which may contribute to morbidity and mortality. We describe endotracheal aspirate samples from seven patients with SARS-CoV-2 pneumonia requiring mechanical ventilation. We note SARS-CoV-2 virions within lower respiratory tract myeloid cells shown by electron tomography, immunofluorescence confocal imaging, and immuno-electron microscopy. Endotracheal aspirates are primarily composed of mononuclear and polymorphonuclear leukocytes. These myeloid cells that harbor virus are frequently positive for CD14 and/or CD16 and most display an inflammatory phenotype marked by expression of IL-6 and tissue factor mRNA transcript and protein expression.

## Brief Report

SARS-CoV-2 pneumonia may induce an aberrant immune response with brisk recruitment of myeloid cells into the airspaces.^1–3^ Although the clinical implications are unclear, others have suggested that infiltrating myeloid cells may contribute to morbidity and mortality during SARS-CoV-2 infection.^1,4,5^

We examined endotracheal aspirates (Supplemental methods) from seven patients with severe COVID-19 pneumonia who required mechanical ventilation (Table 1): three patients required extra-corporeal membrane oxygenation and two patients were deceased by 60 days of follow-up from intensive care unit admission. Endotracheal aspirates were primarily composed of mononuclear and polymorphonuclear leukocytes (Figure 1A, range 70.6-97.5% of nucleated cells). Electron tomography (ET) of endotracheal aspirates revealed intra-cellular localization of presumptive SARS-CoV-2 virions in mononuclear leukocytes (representative image, Figure 1B) and polymorphonuclear leukocytes (representative image, Figure 1C). The identification of SARS-CoV-2 virions by ET was consistent with immune-electron microscopy using an antibody against the Nucleocapsid protein of SARS-CoV-2 that confirmed the presence of virus in CD14+ cells in the lower airways (representative image, Figure 1D). Quantitative imaging of ETA cells revealed SARS-CoV-2 Nucleocapsid protein expression (Figure 1E; n=6; Patient 7 did not have sufficient ETA available for imaging), many of which were also positive for CD14 or CD16 immunostaining (Figure 1F; representative image displayed in Figure 1G). Myeloid cells that expressed SARS-CoV-2 Nucleocapsid protein also frequently expressed IL-6 and tissue factor protein (Figure 1H). Finally, we noted that endotracheal aspirate myeloid cells showed *IL6, F3* and *CD14* transcripts (representative image, Figure 1I).

**Table 1:**
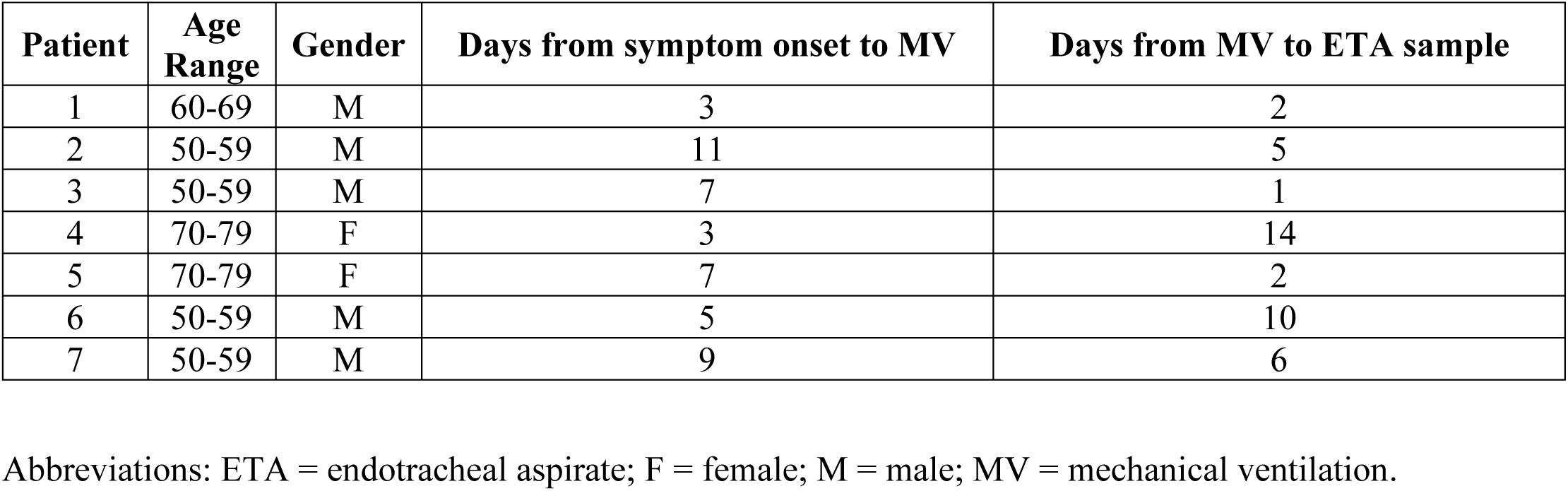
Clinical characteristics of seven patients with severe SARS-CoV-2 pneumonia

**Figure 1.**
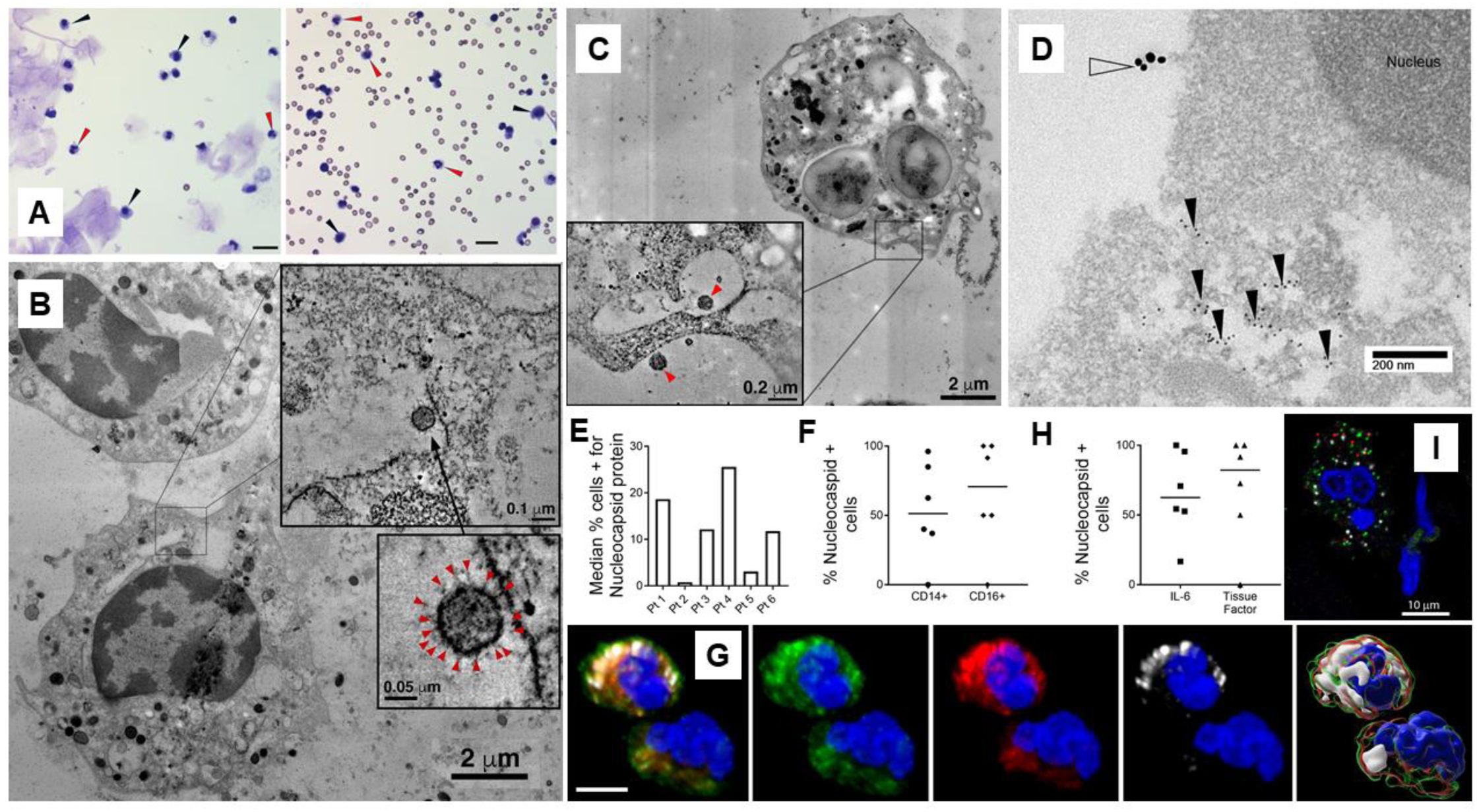
(A) Representative images of cytospins prepared from endotracheal aspirate (ETA) samples – Patient 3 is on the left and Patient 6 is on the right. Black arrowheads denote mononuclear cells and red arrowheads denote polymorphonuclear cells. Black scale bar in lower right portion of each image indicates 20 microns. (B) Electron microscopy (EM) overview of lower respiratory tract mononuclear leukocyte (presumptive macrophage) from Patient 4 with upper inset demonstrating the region indicated by the square that shows tomographic slice with presumptive SARS-CoV-2 virion in a smooth-walled compartment or surface invagination. Lower inset shows higher magnification tomographic view of presumptive virion with apparent spike proteins indicated by red arrowheads. (C) Polymorphonuclear leukocyte (presumptive neutrophil) from Patient 7 with inset showing the region indicated by the square in the overview containing presumptive SARS-CoV-2 virions (red arrowheads). (D) ImmunoTEM of lower respiratory tract mononuclear leukocyte from Patient 6 with CD14 (18 nm gold colloid, open arrowhead) surface immunostaining and internal immunostaining of SARS-CoV-2 Nucleocapsid protein (6 nm gold colloid, black arrowheads at clusters of staining) (E) Quantitative immunofluorescence with median percentage (n=3 slides per patient) of total ETA cells that expressed SARS-CoV-2 Nucleocapsid protein (n=6 patients, Patient 7 did not have sufficient endotracheal aspirate for immunofluorescence staining). (F) Percentage of endotracheal aspirate cells that co-expressed Nucleocapsid protein and CD14 or CD16 (n=6 patients). (G) Representative montage from a single polymorphonuclear cell illustrating co-localization by immunofluorescence. Panels from left to right show: a) merge, b) CD14 (green), c) IL-6 (red), d) SARS-CoV-2 Nucleocapsid protein (white), and e) Imaris (Bitplane) surface rendered image of the overlapping areas of labeling. The blue nuclear stain in all panels is DAPI. White scale bar is 10 microns. (H) Quantitative immunofluorescence of Nucleocapsid protein positive cells that co-expressed IL-6 or tissue factor (TF) (n=6 patients). (I) Representative in situ localization of *CD14* (green), *IL6* (white), and tissue factor or *F3* (red) transcript as well as DAPI nuclear staining (blue) in ETA myeloid cell.

Taken together, our findings suggest that myeloid cells found in endotracheal aspirate samples harbor SARS-CoV-2 and display an inflammatory phenotype marked by expression of IL-6 and tissue factor. The mechanisms by which virions enter lower respiratory tract myeloid cells and survive phagocytic degradation are unclear. However, others have noted human monocyte-derived macrophages to harbor SARS-CoV-1 virus without productive replication *in vitro* and prior reports have shown survival of HIV virions in bone marrow macrophages in a humanized mouse model.^6,7^ The clinical implications of our findings are unknown, but it is intriguing to consider that the purported benefits of dexamethasone in patients with SARS-CoV-2 pneumonia requiring mechanical ventilation^8^ may potentially result from modulation of inflammatory myeloid cells recruited to lung airspaces, which are deleterious in mouse models of SARS-CoV-1 pneumonia.^9^ Further work remains to determine the clinical implications of myeloid cells harboring SARS-CoV-2 virus during severe SARS-CoV-2 pneumonia.

## Data Availability

All relevant data is provided within the manuscript. Requests for additional data should be directed to the corresponding author.

## Supplemental Methods

### Patients

Samples were prospectively collected from patients hospitalized in intensive care units (ICU) at the University of Pittsburgh Medical Center Presbyterian and Shadyside hospitals with SARS-CoV-2 infection as documented by RT-PCR. Patients were enrolled within 72 hours of intubation whenever possible. Upon the informed consent of the patient’s legally authorized representative, patients were enrolled in the University of Pittsburgh Acute Lung Injury Registry and Biospecimen Repository (IRB# PRO10110387), which has been described elsewhere.^10,11^ Longitudinal samples were collected on days 5 and 10 after enrollment.

### Sample collection, handling, and virus inactivation

Endotracheal aspirates were collected from mechanically ventilated patients after pre-oxygenation with 100% oxygen for 2 minutes. 5-10 milliliters of 0.9% sodium chloride solution were instilled via the instillation port on the closed suction system of the endotracheal tube to lavage the airspaces. De-identified samples were placed in biohazard bags and decontaminated per local protocol then transported on ice for further processing in a Bio-Safety level 2+ laboratory space.^12^ Raw endotracheal aspirates were diluted 1:1 in 8% (volume/volume) paraformaldehyde and stored at 4°C overnight to inactivate virus. Following viral inactivation with 4% paraformaldehyde overnight fixation, endotracheal aspirates were washed twice in sterile phosphate buffered saline (PBS) and pelleted at 600 g.

### Electron tomography

The washed cell pellet was re-suspended in 0.5 mL 1% glutaraldehyde then pelleted at 600 g and stored at 4°C. ETA samples were further centrifuged to concentrate the pellet, then transferred to brass planchettes (Ted Pella, Inc.) and ultra-rapidly frozen with a HPM-010 high pressure freezer (Bal-Tec/ABRA, Switzerland). As previously described,^6^ the vitrified samples were transferred under liquid nitrogen to cryotubes (Nunc) containing a frozen solution of 2.5% osmium tetroxide, 0.05% uranyl acetate in acetone. Tubes were loaded into an AFS freeze-substitution machine (Leica Microsystems) and processed at −90°C for 72 hr, warmed over 12 hr to −20°C, held at that temperature for 8 hr, then warmed to 4°C for 1 hr. The fixative was removed and the samples rinsed 4 x with cold acetone, following which they were infiltrated with Epon-Araldite resin (Electron Microscopy Sciences, Port Washington, PA) over 48 hr. Samples were flat-embedded between two Teflon-coated glass microscope slides and the resin polymerized at 60°C for 48 hr. Flat-embedded ETA was observed with a stereo dissecting microscope to ascertain and select well-preserved samples. Suitable regions were extracted with a scalpel and glued to the tips of plastic sectioning stubs. Semi-thick (300-400 nm) serial sections were cut with a UC6 ultramicrotome (Leica Micro-systems) using a diamond knife (Diatome, Ltd. Switzerland). Sections were placed on Formvar-coated copper-rhodium slot grids (Electron Microscopy Sciences) and stained with 3% uranyl acetate and lead citrate. Gold beads (10 nm) were placed on both surfaces of the grid to serve as fiducial markers for subsequent image alignment. Grids were placed in a dual-axis tomography holder (Model 2040, E.A. Fischione Instruments, Export, PA) and imaged with a Tecnai TF30ST-FEG trans-mission electron microscope (300 KeV; ThermoFisher Scientific) equipped with a 2K CCD camera (XP1000; Gatan, Inc, Pleasanton CA). Tomographic tilt-series and large-area montaged over-views were acquired using the SerialEM software package (Mastronarde, 2005). For tomography, samples were tilted + /- 64° and images collected at 1° intervals. The grid was then rotated 90° and a similar series taken about the orthogonal axis. Tomographic data were calculated, analyzed and modeled using the IMOD software package (Kremer et al., 1996; Mastronarde, 2008) on MacPro and iMac Pro computers (Apple, Inc, Cupertino, CA).

### Cytospin preparation and endotracheal aspirate cell differential

Washed cell pellets were re-suspended in sterile PBS prior to spinning at 300 RPM in a Shandon Cytospin3 onto a Superfrost plus microscope slide (Fisher Scientific) then air dried. One cytospin slide was stained with Diff-Quick (Siemens) then evaluated under light microscopy to confirm acceptable spacing and numbers of cells. Cell differentials were performed by manual counts of 200 nucleated cells by three separate authors who were blinded to the others’ results – the mean result of the counts was utilized. Representative images were obtained utilizing an Olympus Provis I fluorescence microscope.

### Immunofluorescence

Cytospin slides were prepared as above then placed in 70% (vol/vol) ethyl alcohol followed by 90% (vol/vol) ethyl alcohol for 10 minutes each, then allowed to air dry. Slides were stored at −20°C for subsequent staining. To perform immunofluorescence staining, cells were washed 3 times in PBS, permeabilized in 0.1% Triton-X-100 in PBS for 20 min, and then washed once with PBS. Cells were then blocked in 20% normal goat serum in PBS for 40 minutes then washed once with 0.5% BSA in PBS (BPBS). Primary antibodies (Supplemental Table 1) were added to cells in BPBS at a 1:100 dilution and incubated the at room temperature for 1 hour. Cells were washed 4 times in BPBS then secondary antibodies at designated dilutions were added for 1 hour at room temperature. Cells were washed 3 times in BPBS then 3 times in PBS. Nuclei were labeled with 0.1% Hoechst’s dye for 30 seconds then rinsed with PBS. Cells were cover slipped using gelvatol (23 g poly(vinyl alcohol) 2000, 50 ml glycerol, 0.1% sodium azide to 100 ml PBS), then viewed on a confocal scanning fluorescence microscope (Nikon A1).

### Pre-embed ImmunoTEM

Cytospins were collected and processed as for immunofluorescence, except primary antibody dilutions were used at 1:50 and incubated overnight at 4°C and secondary antibodies were incubated at a dilution of 1:10 at room temperature for 4 hours. Colloidal gold conjugated secondary antibodies used were goat anti-mouse 18 nm and goat anti-rabbit 6 nm (Jackson ImmunoResearch). After the final wash, samples were fixed in 2.5% glutaraldehyde in PBS for 1 hr. Monolayers were then washed in PBS three times then post-fixed in aqueous 1% osmium tetroxide, 1% Fe_6_CN_3_ for 1 hr. Cells were washed 3 times in PBS then dehydrated through a 30-100% ethanol series then several changes of Polybed 812 embedding resin (Polysciences, Warrington, PA). Cells were embedded by inverting Polybed 812-filled BEEM capsules on top of the cells. Blocks were cured overnight at 37°C, then cured for two days at 65°C. Monolayers were pulled off the slides and ultrathin sections (60-70 nm) of the cells were obtained on a Leica EM UC7 ultramicrotome, post-stained in 4% uranyl acetate for 10 min and 1% lead citrate for 7 min. Sections were viewed on a JEOL JEM 1400 transmission electron microscope (JEOL, Peobody MA) at 80 KV. Images were taken using a side-mount AMT 2k digital camera (Advanced Microscopy Techniques, Danvers, MA).

### RNAscope

Tissue factor (F3), IL-6, and CD14 transcripts were localized to endotracheal aspirate cells by RNAscope Multiplex Fluorescent V2 assay (ACD) according to manufacturer instructions. Briefly, fixed-frozen cytospin sections were incubated with hydrogen peroxide and washed with distilled water. Then slides were placed in 1x target retrieval agent and antigen retrieval was performed at 99°C for 5 minutes in a steamer, then slides were rinsed with distilled water, transferred to Ethanol 100% and dried overnight. The next day, slides were treated with Protease III and immediately treated for RNAscope. CD14 (C1, 520nm), Tissue Factor (C2, 570nm) and IL-6 (C3, 690nm) probes were then hybridized, amplified and developed. Finally, slides were stained with DAPI and mounted with a ProLong Gold Antifade mountant, dried overnight at room temperature in the dark and stored at 4°C until analysis. Slides were viewed and analyzed on a confocal scanning fluorescence microscope (Nikon A1).

### Quantitative Imaging Analysis

Co-expression of Nucleocapsid protein with CD14, CD16, IL-6, and tissue factor was assessed using object-based area overlap analysis (NIS Elements, Nikon Inc., Melville NY). In brief, Z stacks were performed at Nyquist sampling. The data was deconvolved and binary masks for each channel were generated based on emission intensity. Co-expression was assessed using a binary “and” statement was used to determine pixels containing both Capsid protein “and” the marker of interest. Imaris (Bitplane) was used to generate a surface rendered image that illustrated the areas of co-localization in this dataset.

**Supplemental Table 1:** Additional description of antibodies

| **Epitope** | **Species** | **Vendor** | **Catalogue #** | **IF Dilution** | **EM Dilution** |
| --- | --- | --- | --- | --- | --- |
| SARS-CoV-2 Nucleocapsid | Rabbit | Novus | NB100-56576 | 5 µg/ml | 10 µg /ml |
| CD14 | Mouse, Clone: MΦP9 | BD Biosciences | 347490 | 0.25 µg /ml | 0.5 µg /ml |
| CD16 | Mouse, Clone DJ130c | Invitrogen | MA1-84008 | 1 µg /ml | N/A |
| CD142 (TF) | Mouse, Clone HTF-1 | BD Biosciences | 550252 | 5 µg /ml | N/A |
| IL6 | Rat | Novus | NBP2-44953 | 2 µg /ml | N/A |
| IgG | Goat anti-Mouse 18 nm | Jackson ImmunoResearch | 115-215-146 | N/A | 1:10 |
| IgG | Goat anti-Rabbit 6 nm | Jackson ImmunoResearch | 111-195-144 | N/A | 1:10 |
| IgG | Goat-anti-mouse Alexa 488 | Invitrogen | A11029 | 2 µg /ml | N/A |
| IgG | Goat anti-Rabbit Cy3 | Jackson ImmunoResearch | 111-165-003 | 2 µg /ml | N/A |

Abbreviations: IF = immunofluorescence, EM = immuno-electron microscopy

## Acknowledgements

The authors wish to thank the patients and patient families that have enrolled in the University of Pittsburgh Acute Lung Injury Registry. We also thank the physicians, nurses, respiratory therapists and other staff at the University of Pittsburgh Medical Center Presbyterian and Shadyside Hospital intensive care units for assistance with coordination and collection of endotracheal aspirate samples. We thank Nicole Bensen and Caitlin Schaefer at the University of Pittsburgh for assistance with identifying and consenting patients and their families and for assistance with collection of endotracheal aspirate samples. We thank Heather Michael and Lauren Furguiele at the University of Pittsburgh for assistance with processing endotracheal aspirate samples.

This work was supported by Career Development Award Number IK2 BX004886 from the United States Department of Veterans Affairs Biomedical Laboratory R&D (BLRD) Service (W.B.); the National Heart, Lung, And Blood Institute of the National Institutes of Health under Award Numbers K23 HL129987 (G.D.K); P01HL114453 (J.S.L., B.J.M.); and R01 HL136143, R01 HL142084, K24 HL143285 (J.S.L.). Electron and confocal microscopy at the University of Pittsburgh Center for Biologic Imaging was supported by National Institutes of Health Office of the Director awards S10OD010625 and S10OD019973 (S.C.W.). Electron tomography at the California Institute of Technology was supported by a George Mason University Fast Grant (P.J.B.), National Institute of Allergy and Infectious Diseases (NIAID) Grant 2 P50 AI150464 (P.J.B.), and that EM was performed using a TF-30 electron microscope maintained by the California Institute of Technology Kavli Nanoscience Institute. The content is solely the responsibility of the authors and does not necessarily represent the official views of the National Institutes of Health, Department of Veterans Affairs, or any other sponsoring agency.

